# Extracting meaningful features from time series analysis of plasma analytes in clinical research

**DOI:** 10.64898/2026.10.08.26365091

**Authors:** Paul S. Blank, Adriana E. Golding, Ivonne Morales, Hang Waters, Elena Mekhedov, Ludmila Bezrukov, Rebecca D. Wachter, Irina Mikhailenko, Robert H. Brown, Carsten G. Bönnemann, Andrew P. Demidowich, Minal S. Jain, Jack A. Yanovski, Joshua Zimmerberg

**Author notes:** Corresponding author: Joshua Zimmerberg. Division of Infectious Diseases and Tropical Medicine, Center of Infectious Diseases, Heidelberg University Hospital, Heidelberg, Germany (Ivonne Morales).

## Abstract

While absolute blood biomarkers have long guided diagnosis, their rate of change often provides greater diagnostic value. For the Limb Girdle Muscular Dystrophies studied here, increased muscle weakness is associated with abnormal sarcolemma repair. Time course analyses were made to changes in muscle-derived plasma enzymes after modest physical activity of ambulatory inpatients. Blood samples at 20 selected time points at variable intervals were obtained at rest, and before and after routine morning activities and motor function testing. After these activities, plasma enzyme levels increased in a similar way, suggesting damage and leakage from skeletal muscle. The levels then decreased in a similar way, suggesting that muscle cells stopped leaking and enzymes were subsequently cleared from the blood. The complex temporal changes were modeled using both continuous time and data features to reduce the number of parameters required to describe the irregularly sampled, interrupted, time series data. Unlike individuals with cardiac muscle ischemia or following intense exercise, there was no extended post-injury rising phase of enzyme levels. In contradistinction, levels fell once activity ceased. Time series analyses of blood biomarkers most likely provide an assay and index of sarcolemma fragility and by extension skeletal muscle damage.

## Introduction

The use of blood analytes is a longstanding practice in diagnosing and monitoring the clinical course of disease. Notably, it is often the longitudinal rate of change of analytes, commonly referred to as “biomarkers” ^1^, that holds greater significance than single, absolute values. The changes over time in glucose concentrations after a meal and the variations of cardiac creatine kinase (CK-MB) in plasma after cardiac damage are examples of dynamic biomarkers that play crucial roles in the diagnosis and monitoring of specific health conditions, e.g. diabetes mellitus and acute myocardial infarction (AMI) ^2–4^. Time course analysis, requiring timely quantification of biomarkers, drove the development of newer lab assays ^5^. Newer candidate biomarkers with better sensitivity, selectivity, or affordability are discovered via their kinetic similarities: the time course of CK-MB release after AMI was the gold standard that informed myoglobin and troponin-C analyte research. Importantly, it is the *time course* of troponin C release from cardiac muscle into plasma that improves AMI diagnosis and is currently the standard of care.

Reasoning by analogy that a loss of barrier functions of the cardiac muscle membrane leads to cardiac enzyme elevation in blood, and mindful of many studies showing CK release in vigorous exercise, this study explores if features of the *time course* of non-vigorous activity-dependent release of CK and other biomarkers from skeletal muscles in individuals suffering from certain muscular dystrophies (MD) would *itself* be a biomarker. Unlike myocardial infarction or heat exhaustion-related rhabdomyolysis, there is no identified acute single episode of skeletal muscle injury in individuals suffering from limb girdle muscular dystrophies (LGMD). Rather, due to the mutated proteins, affected skeletal muscle sustains ongoing microscopic injury during activities of daily living. Every muscle contraction generates mechanical stress on the muscle fiber membrane, small sarcolemma tears, and incompletely repaired injury, resulting in chronic progressive myofiber degeneration and elevated CK levels prior to sustained muscle loss.

Here, initial findings from a pilot study to assess biomarkers of changes in barrier function of skeletal muscle in individuals with LGMD are reported. The hypothesis was that there would be significant and reproducible activity-dependent changes in plasma protein levels on short timescales (minutes to days) in affected individuals, and that these changes could serve as biomarkers for sarcolemma fragility. To test this hypothesis, a short-term investigation with multiple blood draws before and after defined levels of physical activity was conducted. There were significant activity-dependent changes in plasma alanine and aspartate aminotransferase (ALT, AST), CK, and lactate dehydrogenase (LDH). Furthermore, time-series analyses identified three of the four enzymes (ALT, AST, CK) as having similar and reproducible rates of clearance. Time-series analyses of plasma ALT, AST, and CK before and after activity may serve as sensitive and quantifiable biomarkers of sarcolemma fragility.

## MATERIALS AND METHODS

### Standard Protocol Approvals, Registrations, and Patient Consents

Participants in an ongoing pilot study (ClinicalTrials.gov Identifier NCT01851447) to assess changes in individuals clinically affected with genetically confirmed muscular dystrophy were recruited through the Jain Foundation, Muscular Dystrophy Association, and patient/parent support groups. Individuals taking anti-inflammatory medications or having other co-morbidities were excluded as these could be study confounders. The protocol was approved by the NIH Clinical Center and NICHD Institutional Review Board and all participants provided written informed consent for their participation.

### Study Procedures

All participants were admitted to the NIH Clinical Center for a 5-day inpatient stay. Upon admission, an intravenous (IV) catheter was placed in the arm, and all subsequent blood draws were obtained from this IV or a replacement. Tourniquets were not used to assist with blood draws as they could falsely affect measured enzyme concentrations. On the morning of Day 1, participant health was assessed with baseline labs: Complete Blood Count (CBC), Prothrombin Time (PT), Partial Thromboplastin Time (PTT), Creatinine level, Basic Metabolic Panel, Hepatic Panel, Cystatin C, high-sensitivity C-reactive protein (hsCRP), Hemoglobin A1C, Thyroid Stimulating Hormone, and Free T4 were measured using standard procedures by the Clinical Center Department of Laboratory Medicine.

A total of 20 blood samples were obtained over the 5-day inpatient period (Figure 1); serial blood draws were performed according to the following schedule. Day 1: #1, beginning of bedrest at night; Day 2: #2, 3 AM in supine position, #3, 7 AM after bedrest but prior to any activity in supine position, #4, 30 min following routine morning activity (e.g., toilet, shower, dressing, breakfast), #5, immediately after physical therapy assessment and every 5 min thereafter for 30 min, #6-11, and 2, 4, 6 hours post-assessment #12, 13, 14; Days 3-5: #15, 7 AM before routine morning activity and #16, 30 min following routine morning activity, #17, 18 on day 4, #19, 20 on day 5. ALT, AST, CK, and LDH were measured using colorimetric assays on Roche Cobas analyzers with lower limits of analytical measurement range of 5, 5, 10, and 7 U/L and imprecision (Total CV; %) less than or equal to 3.3, 2.3, 2.7 and 3.3, respectively.

**Figure 1:**
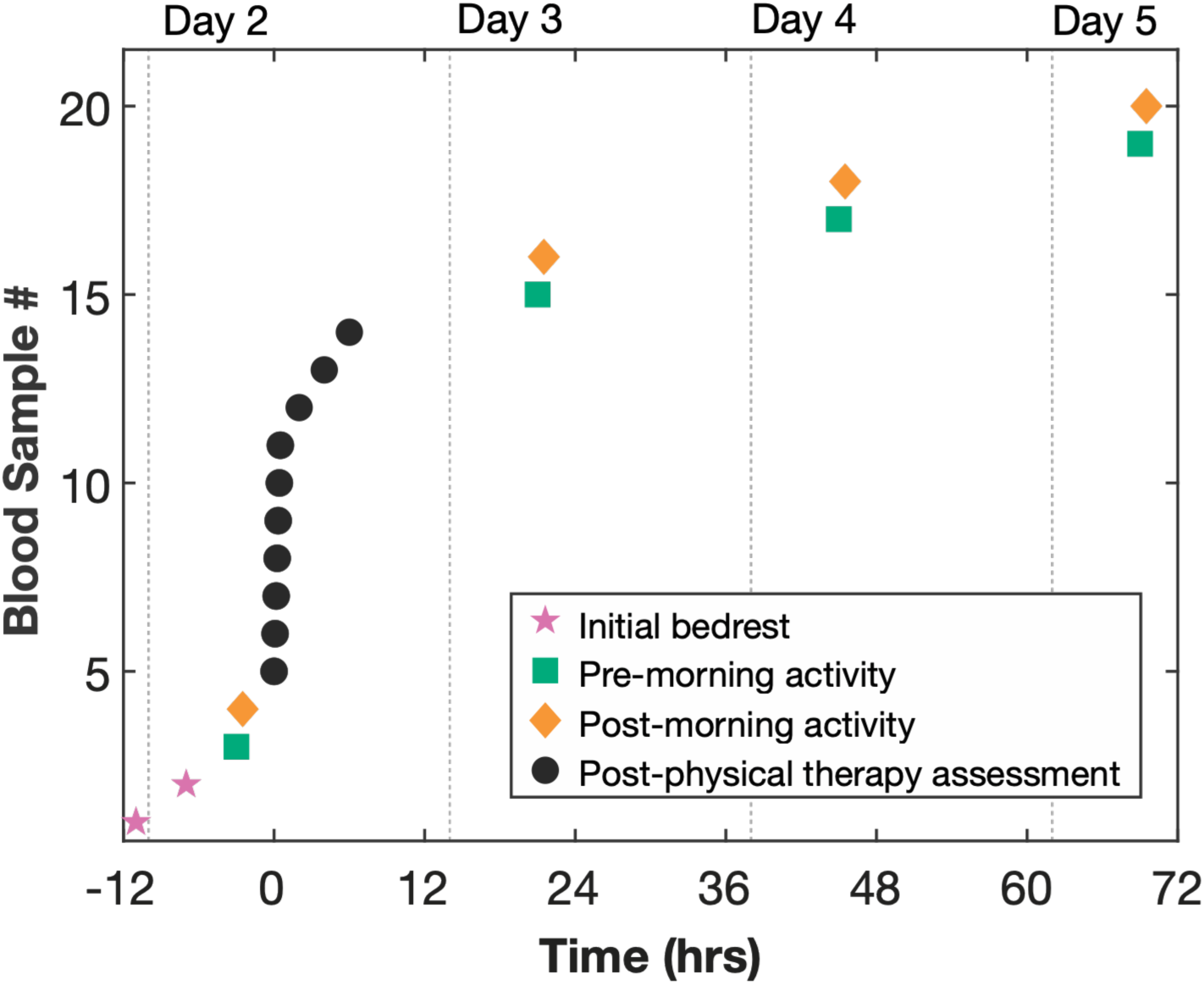
Phlebotomy protocol. Schematic of blood draw schedule over the course of 5 days. Blood was drawn at (1) beginning of bedrest on Day 1, (2) during bedrest on Day 2 at 3AM (pink stars); (3, 15, 17, 19) daily at 7AM after bedrest but prior to any activity in supine position (green squares); (4, 16, 18, 20) 30 min after routine morning activity each day (orange diamonds); (5-14) at high frequency over a short time interval following physical therapy assessment (black circles; time 0 on x-axis, every 5 min for draws 6-11, 90 min between draws 11 and 12, and 120 min between draws 12, 13 and 14).

### Data Analysis

Log-normal Distributions: Large population variability precluded simple averaging strategies. Across all participants, enzyme activity concentrations were better described by a log-normal distribution than a normal distribution for each analyte, as confirmed by comparing Akaike Information Criterion (AIC) values for both model fits. The log-normal model’s AIC was lower than the normal model’s AIC by ΔAIC = 94.14 (ALT), 77.38 (AST), 92.26 (CK), and 46.49 (LDH). Following standard AIC-based model-selection guidance ^6^, a ΔAIC exceeding 10 indicates essentially no empirical support remains for the higher-AIC (normal) model relative to the lower-AIC (log-normal) model — so the log-normal distribution was adopted for all subsequent analyses. The enzyme measurements were not biased at low levels by the assay’s limits of detection or quantification. Consequently, all enzyme activity concentrations were log transformed prior to subsequent data analysis. Log transformation also aids in identifying a single exponential process or sums of exponential processes with sufficiently different time constants via the number of identified linear segments. When applied to time-dependent compartmental analysis, log transformation of exponential processes results in one or more linear segments. Additionally, the data from two individuals with the same exponential decay process but different amplitudes will be represented by two parallel lines following log transformation. In theory, multiple individuals could be compared through the weighted average of their individual parameters sets.

Z-score Transformation: In practice the results obtained in this study were too sparse to be fit by more complex models. To further reduce variability due to differences in individual participant activities, individual Z-score transformation was used, where Z(t_i_) = (Value(t_i_)-m)/ s and i = 1-20 (m = individual’s average activity and s = standard deviation (SD) over all 20 samples per individual). Z-score transformation facilitated comparisons of temporal features and structures between data sequences with different amplitudes; the average of individual Z-scores well described the behavior of the entire participant cohort. The slope of the log transformed data can be recovered by dividing the slopes in the Z-score model by the average SD of the cohort data set provided that this parameter is well defined, as it was for our data. Decay times appropriate for time dependent compartment models were derived using this approach. Note, the Z-score averages are no longer necessarily Z-distributed but can be renormalized to create a new Z distributed parameter. For group comparisons (t-Test or ANOVA), variance homogeneity was evaluated using Levene’s statistic (absolute) and when heteroscedastic, Welch’s test was used for group comparisons.

Two models describe the Z-score averages. The first, designated a feature model, captured defined data set characteristics using global fitting and constraints on specific time series components. The second, designated a consensus, continuous time series model, used the decay constants derived from the feature model to describe the expected time dependence of a continuous time series. This model may be useful in developing experimental sampling strategies based on the specific features of interest.

### Feature and continuous time models

A) Feature model: *I_1_*, *I_2_*, and *I_3_* are linear intercepts, the linear slopes τ_1_, τ_2_, and τ_long_, correspond to exponential decay times, t is the time relative to draw 5, δ is 1/0 for a 7:30/7:00 AM blood draw and max(0,x) is a hinge function (0 when x<0, x otherwise; B) Continuous time model: amplitudes Amp_1_ and Amp_2_, ΔZ, and fraction f_1_ are fitting parameters and all the τ’s were fixed at the values obtained using the feature model. The fractions f_1_ and (1-f_1_) represent the relative contribution of the processes with exponential decay τ_1_ and τ_2_, respectively.

A. Feature Model:

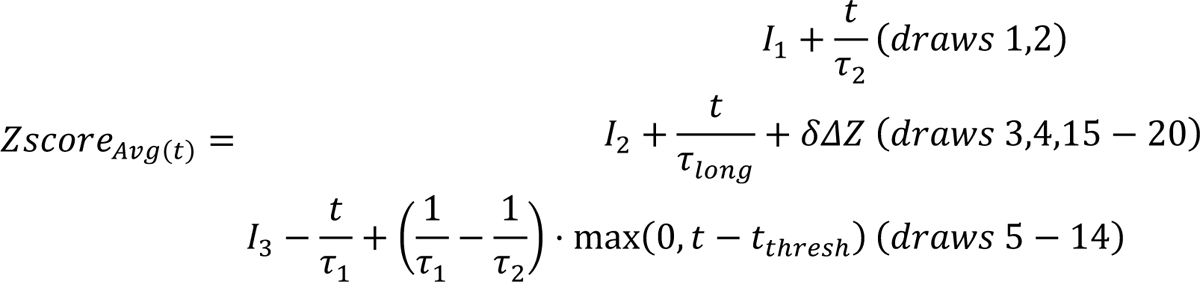
B. Continuous Time Model:

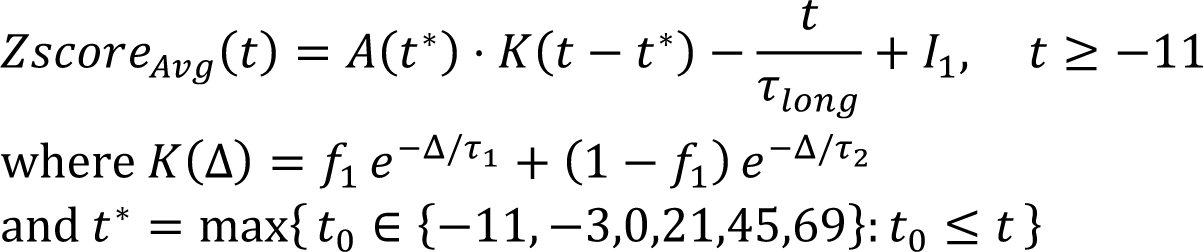

The consensus time series was created by averaging ALT, AST, and CK data for all 11 participants, with subsequent correlation analysis to remove poorly correlated participant data ^7^. Four individual time series of the original 33 were dropped and a new average and variance were calculated from the remaining 29 time series. This average was fit by the continuous time series model with fixed decay times and established by the feature model, where amplitudes (2 parameters – Amp1 and Amp2), decay fractions in the high frequency region (1 parameter-f1) and ΔZ-score (1 parameter) were evaluated by the fitting (normalized inverse variance weighting).

The expected data set is 220 per analyte, or 880 total. However, a total of 2 blood draws were not collected (8 missing values) and 3 outliers (identified using MATLAB outlier detection) were dropped leading to 869 data points analyzed. Data analysis was conducted using Excel v16.107.3 (Microsoft Corp., Redmond, WA) and MATLAB r2025b (The MathWorks, Inc., Natick, MA). Errors are reported as 95% confidence intervals or standard error of the mean (SEM). Figures were created using the data visualization toolbox Gramm (https://doi.org/10.21105/joss.00568).

## RESULTS

A total of 11 participants were enrolled. The cohort consisted of five (four females, one male) LGMD Type 2B/R2 Dysferlin-related (R2), five (three females, two males) LGMD Type 2L/R12 Anoctamin-5-related (R12), and one (one male) LGMD Type 2I/ R9 FKRP-related (R9) individuals. All participants exhibited normal kidney function (eGFR range 72 to >120 mL/min/1.73 m^2^). Participants with LGMD R2 were younger than those with LGMD R12 (36.7 +/- 5.3 or 32.5 (23.8-55.1) and 54.1 +/- 1.6 or 55.5 (48.5-57.5); mean +/- SEM or median (range), n=5, p = 0.027 unequal variance t-test or p = 0.028 Kruskal-Wallis test). There were no differences in the average height and average BMI of the R2 and R12 participants (equal variance t-test or Kruskal-Wallis test). Participants self-identified Race (64% White, 18% African American, 9% Asian, and 9% Multiracial) and ethnicity (91% Not Hispanic or Latino, 9% Hispanic or Latino).

Plasma ALT, AST, CK, and LDH enzyme activity concentrations varied between participants and between LGMD types (R2, R9, R12) (Figure 2A, Additional Figure 1). Following log transformation, when grouped separately, or together (R2, R12, R2 and R12, and R2, R9, and R12), only ALT (all groups), AST (R2 only), and LDH (R2 only) showed equal variance. ALT, AST, CK, and LDH were lower in R12 compared to R2, over all samples (mean(log(Analyte_R12_))-mean(log(Analyte_R2_)) = −1.01 (−1.14, −0.90), −0.59 (−0.70, −0.50), −1.62 (−1.78, −1.47), −0.58 (−0.67, −0.50), respectively; mean differences (99% lower, upper bootstrap CI, 10^6^ resamples). Mean values within each group were unequal (ANOVA analysis; p<10^-10^). Consequently, for each analyte, direct averaging across all participants resulted in large uncertainty (95% bootstrap confidence interval; 5,000 resamples) around the average (Figure 2B). Large uncertainties exist even when averaging by LGMD type.

**Figure 2:**
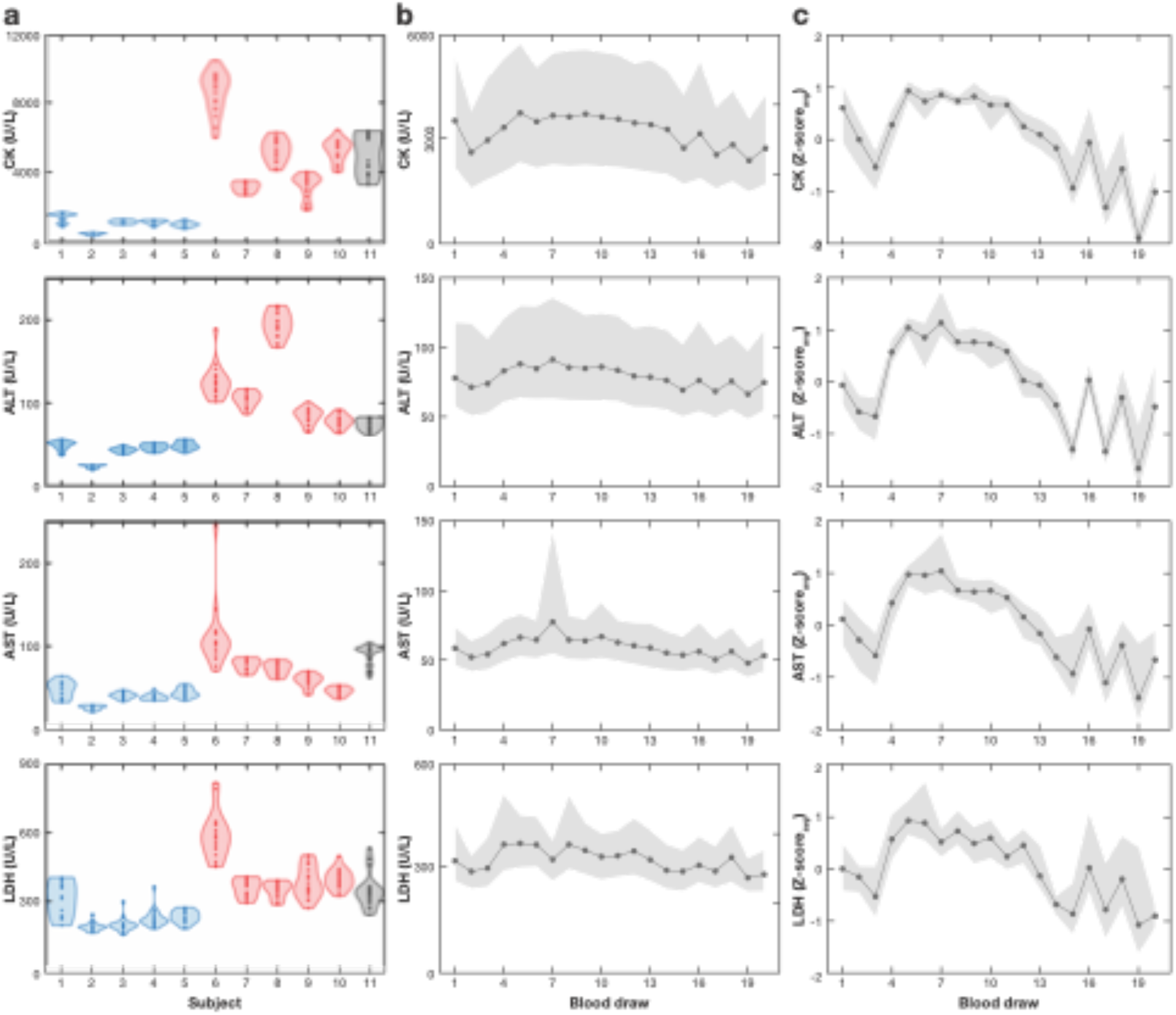
Significant variability in plasma enzymes between participants requires meaningful transformation for time-series analysis of entire cohort averages. **a)** Violin plots of plasma ALT, AST, CK, and LDH by participant, clustered by LGMD subtype (blue-R12 Anoctamin5-related; red-R2 Dysferlin-related; black-R9 FKRP-related). b) Average over all participants (solid line) and 95% bootstrap confidence interval (shading) for each enzyme by blood draw. c) Average over all participants (solid line) and 95% bootstrap confidence interval (shading) of Z-transformed plasma enzyme by blood draw. The Z-score_avg_ represents the average of all participants’ Z_i_ values calculated using a participant’s plasma enzyme average (m) and SD (s), where Z = (Draw_i_-m)/ s; i = blood draws 1-20, and Z_avg_ = <Z_i_>_j_ where j = participants 1-11).

The differences between participants (variance and mean) within groups and between groups (inter- and intra-personal variation) highlights the data complexity and the requirement for feature-preserving analysis. When each participant’s time course for any one analyte was examined (Additional Figure 1), similar responses for each analyte were found: decreasing enzyme levels with rest and increasing enzyme levels with activity. To both examine correlations between LGMD R2 and R12 populations and the population statistics, large variations between individuals were handled using Z-score transformation. First, each participants’ log transformed data underwent a Z-score transformation, whereby we report the difference between each log data value and the participant’s own average over all 20 blood draws normalized by the standard deviation (Additional Figure 1). This Z-score transformation was then averaged (Z-score_avg_) over all participants for each blood draw (Figure 2c). The averaging of the Z-transformed data resulted in reduced uncertainty (95% bootstrap confidence interval; 5,000 resamples) around the average. The trends observed in the Z-score_avg_ reflect those of each individual participants’ Z-scores over the course of the visit (Figure 2, Additional Figure 1).

Significant changes in enzyme activity were detected following 30 minutes of routine morning physical activity (Figure 3). The ΔZ-score change in ALT, AST, CK, and LDH across all participants between pre- and post-morning activity were (1.17±0.14; n=43), AST (0.78±0.24; n=43), CK (0.82±0.12; n=43), and LDH (0.61±0.38; n=42 (ΔZ-score +/- 95% CI; n)), respectively. The Z-score_avg_ for each enzyme had similar temporal features (Figure 4). The magnitudes of the activity-correlated increases were approximately the same for all enzymes. The LDH Z-score_avg_ 95% confidence intervals were greater than the other enzymes, which is consistent with additional differences observed between AST, ALT, CK and LDH described below.

**Figure 3:**
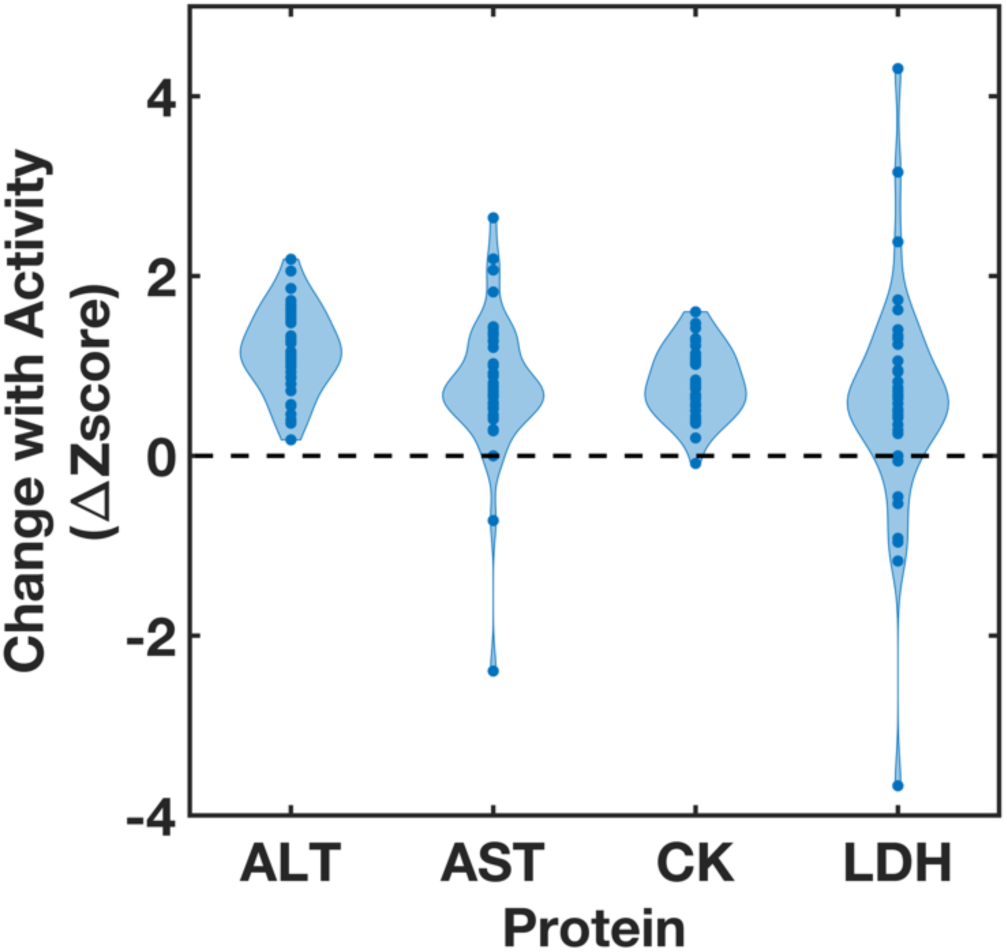
Enzyme activity significantly increases following physical activity. Significant changes in CK, ALT, AST, and LDH enzyme activity (change with activity) are detected following 30 minutes of routine morning physical activity (ΔZ score) across all participants.

**Figure 4:**
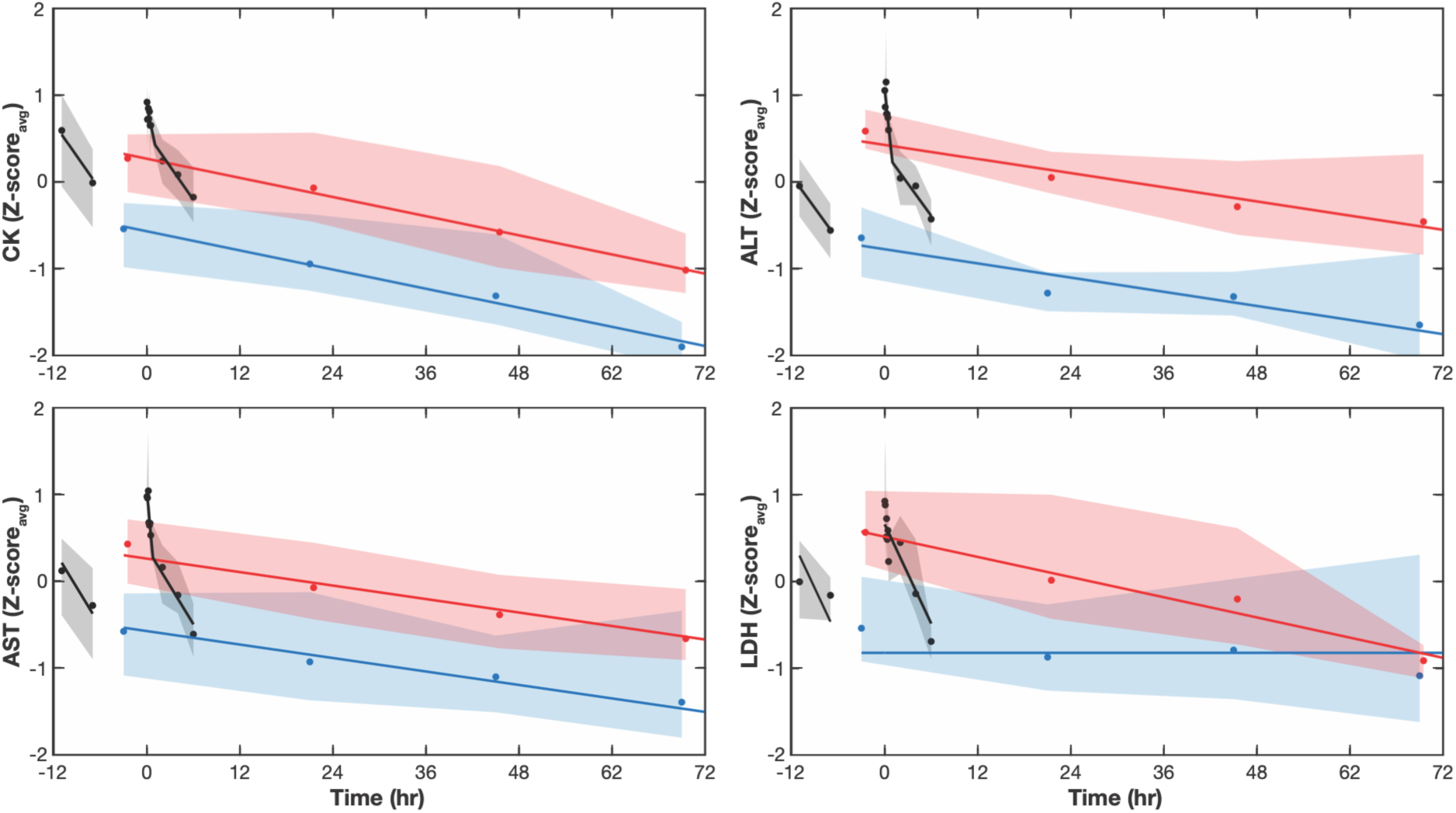
Average plasma enzyme Z-scores reveal three distinct kinetic trends. Average (line) and 95% bootstrap confidence interval (shading) of participant plasma ALT, AST, CK, and LDH Z-scores plotted against time (in hours) with physical assessment at time 0. Blue and red displays correspond to pre- and post-morning activity, respectively. Black displays correspond to post-arrival and post-physical assessment. Blue, red, and black displays are associated with similar decreasing, weeklong trends that are uniformly shifted; black is a relatively faster decay process.

The largest increases in plasma ALT, AST, CK, and LDH occurred following routine physical therapy assessment (Day 2) typically given during clinic visits (Figures 2, 4). Upon arrival at the Clinical Center, plasma enzymes were elevated, perhaps due to high levels of exertion associated with travel. Arrival enzyme levels were comparable to post-assessment levels, as indicated by ratios ∼ 1 for arrival to the post-assessment values (Sample 1: Sample 5). The average ratios for ALT, AST, CK, and LDH, are 0.90 (+/- 0.04), 0.89 (+/- 0.06), 0.96 (+/- 0.08), and 0.87 (+/- 0.11), (mean +/- 95% CI, n = 11), respectively. The gradual decline in plasma enzymes over the following four days may be a result of overall decreased activity in the Clinical Center, as bedrest and reduced physical activity are known to be correlated with reduced plasma enzyme concentrations ^8^.

Other characteristic features observed in the average plasma ALT, AST, and CK Z-score values include (a) elevated enzyme activities on arrival that decreased overnight; (b) two decay components during the high frequency data collection following assessment; (c) a longer decay trend observed over the week that tracks the supine, pre-morning activity blood draws (samples 3, 15, 17, 19), and (d) a similar decay trend observed with post-morning activity blood draws, offset by a constant at each time point (samples 4, 16, 18, 20).

### Analysis of data with a new feature model

The similarities and differences between analytes were then evaluated using global fitting of a feature model (Figure 4). A description of this evaluation follows. (a) The longest decay trends are modeled as parallel offset lines, corresponding to single exponential decay processes modified by a constant offset (ΔZ-score). This offset represents the change in Z-score_avg_ that correlated with morning activity. (b) The high-frequency data are modeled as two piecewise, continuous lines corresponding to two exponential decay processes. (c) The initial, overnight decrease in Z-score was best characterized by the larger of the two exponential decay values, evaluated from the high frequency data and described above in (b). Values and fitting metrics for the feature model parameters are summarized in Table 1.

**Table 1.**
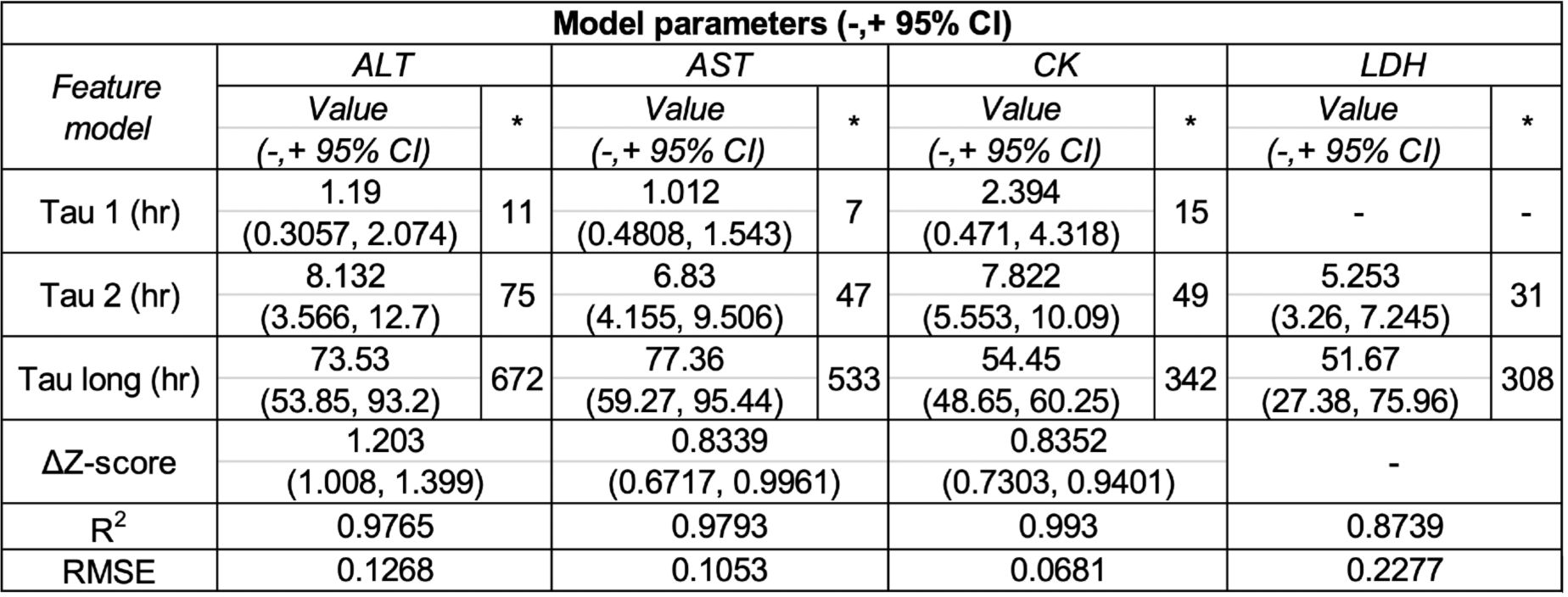
Model parameters for ALT, AST, and CK fit with a feature model consisting of two parallel linear trends and a two-component mixture in the high frequency data region. *Average standard deviation adjusted time constant; R^2^ is adjusted by the number of fitting parameters; root-mean squared error (RMSE). LDH fit with a feature model consisting of a constant background, a linear trend, and a single decay in the high frequency data region.

| Model parameters (-,+ 95% CI) |  |  |  |  |  |  |  |  |
| --- | --- | --- | --- | --- | --- | --- | --- | --- |
| Feature model | ALT |  | AST |  | CK |  | LDH |  |
|  | Value | * | Value | * | Value | * | Value | * |
|  | (-,+ 95% CI) |  | (-,+ 95% CI) |  | (-,+ 95% CI) |  | (-,+ 95% CI) |  |
| Tau 1 (hr) | 1.19<br>(0.3057, 2.074) | 11 | 1.012<br>(0.4808, 1.543) | 7 | 2.394<br>(0.471, 4.318) | 15 | - | - |
| Tau 2 (hr) | 8.132<br>(3.566, 12.7) | 75 | 6.83<br>(4.155, 9.506) | 47 | 7.822<br>(5.553, 10.09) | 49 | 5.253<br>(3.26, 7.245) | 31 |
| Tau long (hr) | 73.53<br>(53.85, 93.2) | 672 | 77.36<br>(59.27, 95.44) | 533 | 54.45<br>(48.65, 60.25) | 342 | 51.67<br>(27.38, 75.96) | 308 |
| ΔZ-score | 1.203<br>(1.008, 1.399) |  | 0.8339<br>(0.6717, 0.9961) |  | 0.8352<br>(0.7303, 0.9401) |  | - |  |
| R<sup>2</sup> | 0.9765 |  | 0.9793 |  | 0.993 |  | 0.8739 |  |
| RMSE | 0.1268 |  | 0.1053 |  | 0.0681 |  | 0.2277 |  |

While similar, there were noteworthy features in the LDH Z-score values that differed from the other analytes. LDH Z-scores could not be modeled with the same feature set but required a different parameterization where the offset varied with time leading to a different decay trend and the long, supine, pre-morning activity trend was absent and best described by a constant value. It is an important distinction that LDH is best described by a different feature model.

Log transformation of an exponentially decaying function results in a linear function; upon averaging the transformed Z-scores across the entire cohort, the resulting decay times were weighted by the standard deviation (SD) of each individual participants’ data. The time constants obtained by fitting the averaged transformed Z-score data were converted, on average, to elapsed time thus unperturbed by the scaling of the slope. This was done after establishing that the SD distributions have well-defined averages and variances and adjusting the decay constants accordingly (Figure 3). These decay constants can be used in compartment modeling, and for comparison with reported clearance rates.

The feature model for the log transformed analyte data was simplified, as the two high frequency time constants were not resolvable. This illustrates an additional advantage for the Z-score treatment: less noise leads to better resolution of multiple decays. To compare the Z-score analyses with the log data fits, the two high frequency constants (τ_1_ and τ_2_) were averaged to obtain an approximate equivalent decay for the single high frequency decay in the log data fits (Additional Table 1).

The similarities between the fitting parameters of the feature model for plasma ALT, AST, and CK were consistent with a common underlying temporal model. This conjecture is supported by Pearson correlation analysis between the different analytes which revealed high and significant correlations between analytes (Additional Figure 3). A consensus time series was then developed using the combined plasma ALT, AST, and CK time series data, following removal of minimally correlated time series (4 of 33 with correlation 95% lower limits less than 0; see methods). The consensus time series is described by a 4-parameter continuous time series model capturing the predicted temporal evolution of an analyte described by the model features (kinetic decay constants) identified for plasma ALT, AST, and CK (Figure 5).

**Figure 5:**
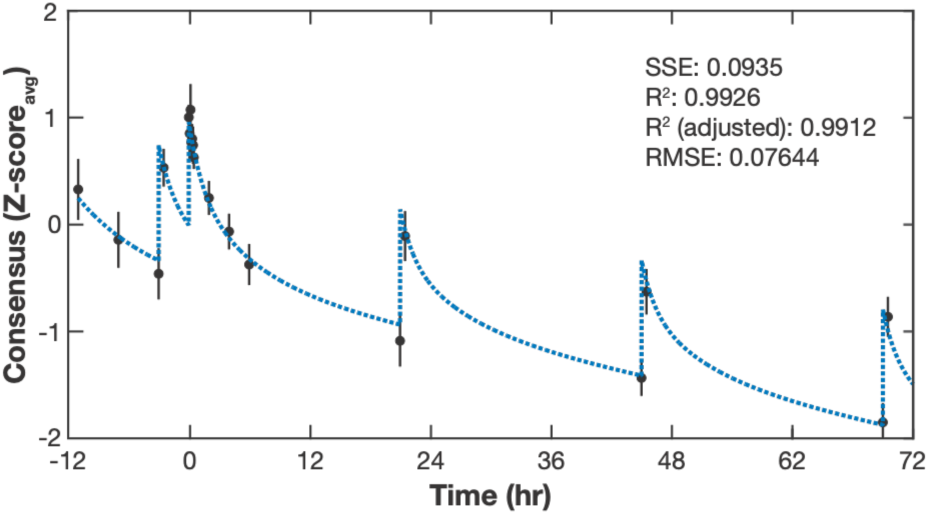
Fitting of cohort consensus data with a continuous time model. The data from a consensus dataset including plasma ALT, AST, and CK (but not LDH) was combined and used in developing a continuous time model. The rates determined from the linear slopes in the feature model were fixed in the continuous time model. The resulting three-parameter fit of the combined data resulted in the following fitted values (-/+ 95% confidence interval): post-assessment amplitudes 0.6039 (0.384, 0.8237) and 1.527 (1.457, 1.597), delta 1.092 (0.9544, 1.21), decay fraction 0.4099 (0.262, 0.5578).

To test the hypothesis that the temporal sequence of measurements is the origin of the significant changes observed in the features model, the analytes measured for each participant were randomly permuted in time. Bootstrap distributions and statistics (average and 95% bootstrap CI) were generated at each time point for all participant sequences with and without permutation (Additional Figure 3). All trends were removed following randomization of the time series data, indicating that model features are dependent upon the temporal ordering of the sequence. This result reinforces the conclusion that the following significant changes observed in the features model reflect the data: 1. Correlations between physical assessment and morning activity, 2. kinetic decay parameters, and 3. morning activity dependent offset (ΔZ-score).

To explore the utility of temporal Z-score analyses in improving experimental design for interventional and longitudinal studies, a power analysis at Power = 0.80 was performed based on the statistical properties of the observed data. The goal was to estimate the number of samples required to identify 100% and 50% changes in ΔZ-score when evaluated from two sequences of blood collected from an individual before and after a daily intervention, using three different probabilities of Type 1 error (α = 0.001, 0.01, and 0.05). Statistical significance was achievable in approximately one month in this example. If this study’s participant cohort is representative of the population of affected individuals, a 50% decrease in ΔZ-score may be detected in 15 – 36 days.

## DISCUSSION

While a single yearly measurement of any dynamically variable biomarker grossly under samples daily changes and obscures yearly trends, this study demonstrates that for our cohort, appropriate time course analysis of the kinetics of highly dynamic plasma ALT, AST, and CK can reproducibly estimate trends in sarcolemma fragility. This allowed a test of the hypothesis that there would be significant and reproducible activity-dependent changes in plasma protein concentrations on short timescales (minutes to days) in affected individuals, and that these changes could serve as biomarkers for sarcolemma fragility. The results were consistent with the above hypothesis.

Using the feature model to analyze enzyme levels over time showed that CK, ALT, and AST were related to physical activity. This relationship was seen across different subjects and groups, even though their starting enzyme levels varied considerably. Activity-dependent changes were observed for all four enzymes, but their kinetics were not the same; whereas the feature model-derived lines for ALT, AST, and CK were parallel over time, LDH’s lines converged (Figure 4). Furthermore, the change in Z-score was the same for ALT, AST, and CK, regardless of the baseline (blue) level before morning activity. This reproducibility in Z-score changes was remarkable, given that each participant arrived at the Clinical Center with a different baseline (elevated compared to subsequent measurements) and the choice and intensity of their morning activities were *ad libitum* for four continuous days. In addition, the decay in enzyme concentrations with bedrest each day was also reproducible and similar between participants. Thus, the consensus model generated from this analysis provides a parametrization of plasma ALT, AST, and CK kinetics that can guide future pathophysiological studies of limb girdle and other muscular dystrophies.

One strength of this study is that our analysis permits investigations of activity-dependent correlations with plasma analytes independent of initial scaling and within small populations of individuals. A limitation is that our time series analysis was based on a small number of time points. Longer time series are expected to improve the parameters of the feature model such that they are usable in time series clustering algorithms when comparing multiple plasma analytes. An additional weakness is specificity: neither the isoenzymes of CK, nor the skeletal muscle specific post-translational isoforms of the CK-MM isoenzyme (I, II, and III) were assayed, nor were isoforms of AST (cytosolic, mitochondrial). While there were no clinical indications of acute myocardial infarction or acute brain damage during the Clinical Center stay that could lead to increases in CK-MB or CK-BB, we cannot exclude that damage to organs other than skeletal muscle contributed to the reported findings. Although only 1% of the total CK in skeletal muscle is CK-MB, a much larger mass of skeletal muscle is involved clinically in the physical activities undertaken in this study. So even though the CK-MB isoenzyme predominates in heart tissue, there is a considerably larger reservoir of CK-MB that can move to blood from skeletal muscle ^9^. Future studies with higher time resolution and increased number of plasma analytes may increase the generalizability of these findings. A key feature of adult-onset muscular dystrophies is that these diseases all progress slowly and show a variable progression ^10^, making traditional trial outcomes, such as decreasing the rate of progression, challenging. Methodologies that facilitate a more rapid identification of treatments that attenuate exertional fragility are needed.

In this study, blood was sampled to capture both short (minutes, hours) and long (days) time periods. This timing allowed correlation of enzyme levels with physical activity for both short and long-term kinetics. The high-frequency blood draws were designed to reveal the kinetics following routine physical assessment, while the data at other times were sparsely sampled to assess longer term differences. The clearance rates calculated here for CK are also consistent with other reports, and plasma ALT and AST have previously been shown to scale with activity in muscular dystrophy and heat exhaustion-related rhabdomyolysis ^11–13^. The transformation of the data described here reflects the overall appearance into blood and clearance from blood, which includes release from muscle, diffusion across the basement membrane, transfer to lymphatics, and passage to blood. Based on previous work ^14^, circadian rhythm is unlikely to significantly contribute to the temporal patterns observed. For individuals with fragile sarcolemma, there is consensus that activity causes skeletal muscle enzymes to be released into the plasma, presumably due to skeletal muscle damage, that are subsequently cleared. Because the data analyses presented here are not dependent on our hypothesis or focus, they can also be used to study clearance more deeply.

Unlike CK, ALT, and AST, the response of plasma LDH to the same morning activity decreased throughout the week, despite the average percent release of the four enzymes being similar (see below). This was curious and may be related to enhanced release of LDH early in the week, but not at the end of the week. Perhaps other activities prior to and during Days 1 and 2 (travel and physical assessments) sensitized the LDH response to morning activities for those days. While almost every tissue has LDH, with heart, liver, and red blood cells obvious candidates for extra-skeletal muscle sources, there is no obvious reason for activity-dependent release of LDH from these other tissues. More work is needed to clarify this issue, including a study of the LDH isoenzyme composition. It is worth noting that LDH also failed to follow the same time course of release from cardiac tissue as CK, AST, myoglobin, and Troponin C following an AMI ^15,16^.

Time course analysis of plasma ALT, AST, CK and LDH supports and expands on findings that both inter- and intra-personal variation are important factors in population studies ^17^. By subtracting “baseline” analyte measures and expressing all changes as variations around “baseline” before and after activity (interrupted time series), significant variations in scaling between individuals are removed and correlated features are revealed. These temporal and correlated features would otherwise be lost if raw data were simply averaged because distributional properties and disparate scaling are not addressed.

Finally, time course analysis of a spectrum of human blood analytes that correlate with physical activity may reveal their source; analytes whose concentrations increase with activity may originate from the same tissue or compartment, and analytes with similar decay times may enter the blood via similar mechanisms. Here, all four enzymes exhibit similar significant increases with activity suggesting that they all originate from the same stimulated source, i.e., skeletal muscle. Prior studies have demonstrated that these enzymes may be released by damaged muscle ^18–20^. The mechanism(s) by which the enzymes enter the blood may combine leakage through sarcolemma wounds, exocytosis, extracellular vesicles ^21–23^ and activity-dependent lymph flow ^24,25^.

## CONCLUSIONS

This study introduces an approach to the design of clinical research and the analysis of interrupted time-series data that can harmonize seemingly disparate results. There were significant and reproducible physical activity-dependent changes in plasma enzyme activity concentrations on short timescales (minutes to days) in affected individuals. Most likely, the plasma enzyme biomarkers presented here represented the release and subsequent clearance of skeletal muscle enzymes due to damage to muscle membranes in these subjects with fragile sarcolemma. Time- and physical activity-dependent measurements may aid diagnosis, long-term longitudinal studies of disease progression and efficacy of gene and drug therapies. The temporal properties of plasma ALT, AST, and CK may facilitate identification of biomarkers with similar, or different, kinetics that further elucidate sarcolemma damage and repair processes. When appropriate perturbations can be found, this approach might facilitate clinical studies for other disease entities where small enrollments hamper population studies.

## LIST OF ABBREVIATIONS

ALT: alanine aminotransferase
ANOVA: analysis of variance
AST: aspartate aminotransferase
CBC: complete blood count
CI: confidence interval
CK: creatine kinase
COA: clinical outcome assessment
hsCRP: high-sensitivity C-reactive protein
LDH: lactate dehydrogenase
LGMD: Limb Girdle Muscular Dystrophy
PTNSAA: prothrombin time North Star ambulatory assessment
PTTPT: partial thromboplastin time prothrombin time
SDPTT: standard deviation partial thromboplastin time
SEM SD: standard error of mean standard deviation
SEM: standard error of mean

## Declarations

### Ethics approval and consent to participate

The protocol was approved by the NIH Clinical Center and NICHD Institutional Review Board and all participants provided written informed consent for their participation.

### Consent for publication

Not applicable.

### Availability of data and materials

Anonymized data not published within this article is available by request from the corresponding author.

## Authors’ contributions

1 Conception or design of the work

2 Data acquisition

3 Data analysis

4 Interpretation

5 Creation of new software

6 Drafting the work or substantively revising

7 Oversight of the clinical care of participants

Paul S. Blank: 1, 3, 4, 5, 6

Adriana E. Golding: 3, 4, 6

Ivonne Morales: 1, 2

Hang Waters: 2

Elena Mekhedov: 2

Ludmila Bezrukov: 2

Rebecca D. Wachter: 1, 6

Irina Mikhailenko: 2

Robert H. Brown Jr.: 1, 6

Carsten G. Bönnemann: 1, 6

Andrew P. Demidowich: 2, 6, 7

Minal S. Jain: 2, 4, 6

Jack A. Yanovski: 6, 7

Joshua Zimmerberg: 1, 2, 3, 4, 6, 7

## Data Availability

All data produced in the present work are contained in the manuscript

## Acknowledgements

The authors thank Ms. Irene T. Rozga, R.N., Dr. Gladys Tataw-Ayuketah, PhD, MPH, RN, and the staff of the Metabolic Clinical Research Unit team for their guidance and proficiency. The authors thank the Jain Foundation for their support of previous pre-clinical research that introduced us to the need for identifying surrogate biomarkers. The authors gratefully acknowledge the contributions of the study participants.

## Additional File 1

**Additional Figure 1:**
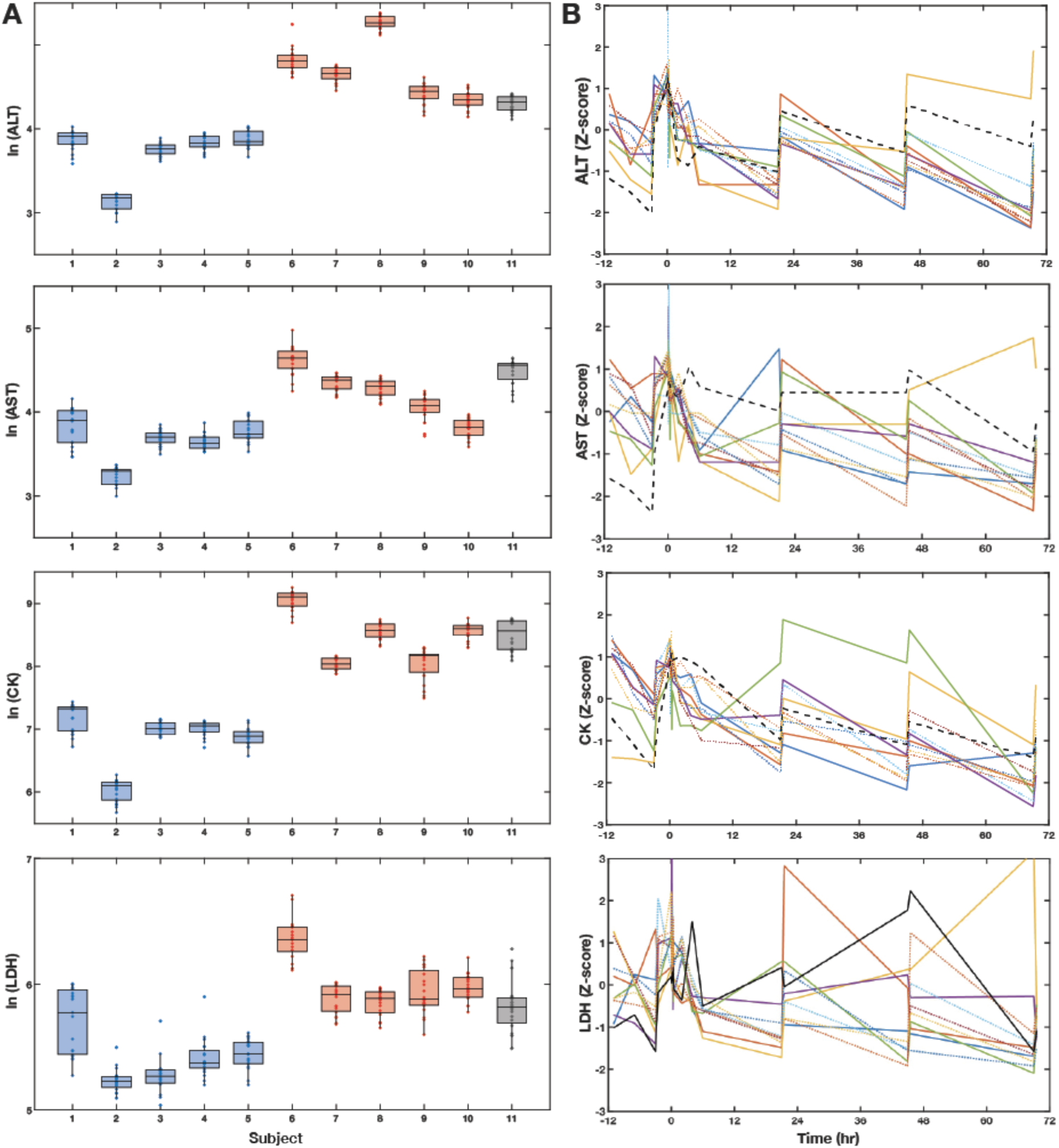
Transformed analysis of plasma ALT, AST, CK and LDH. A) Box plots of log transformed plasma ALT, AST, CK, and LDH by participant clustered by LGMD subtype (blue-R12 Anoctamin5-related; red-R2 Dysferlin-related; black-R9 FKRP-related). B) Z-transformed plasma enzyme levels for each participant over the study time course.

**Additional Figure 2:**
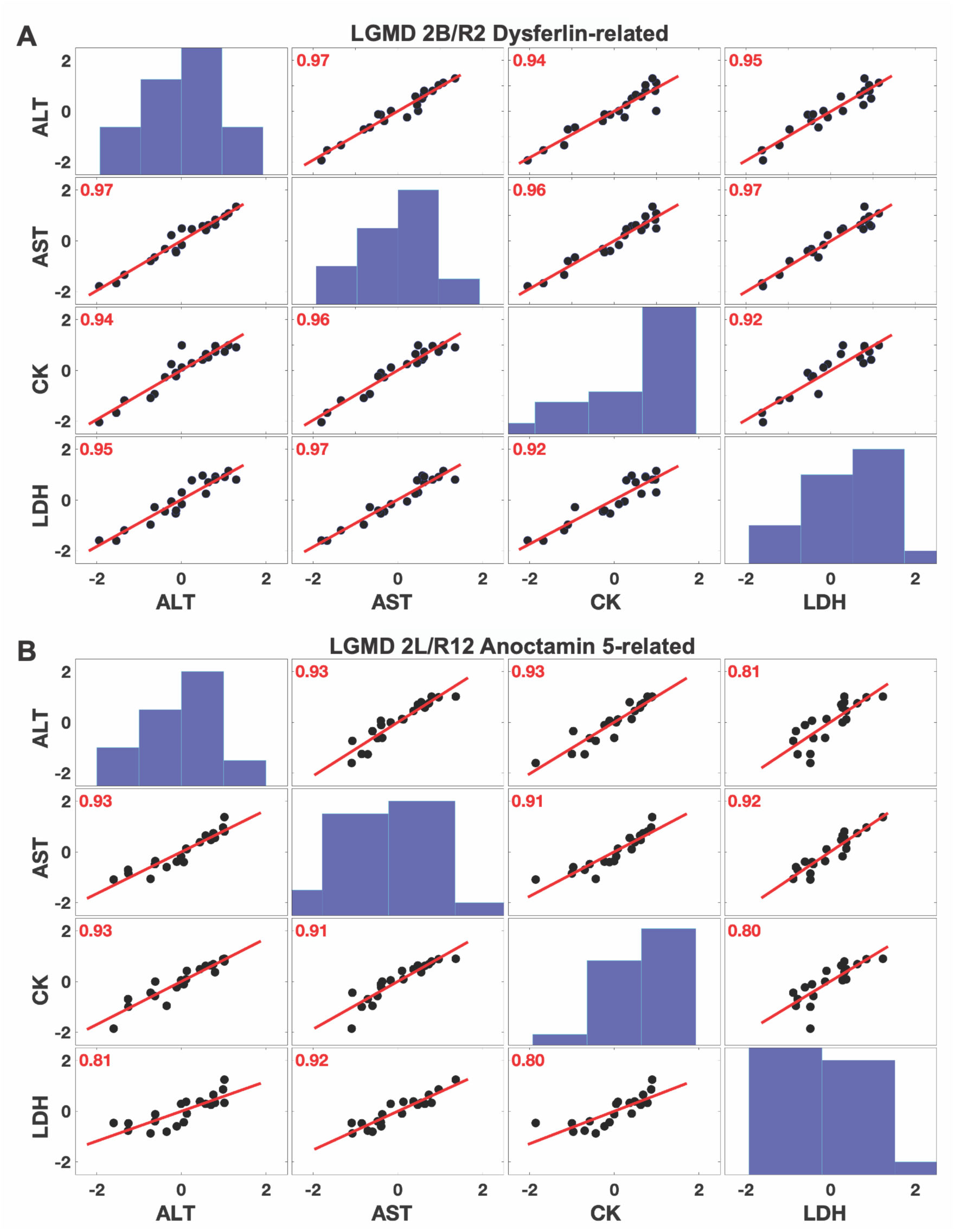
Pearson correlation analysis of ALT, AST, CK and LDH by LGMD subtype. A) LGMD 2B/R2 Dysferlin-related; B) LGMD 2L/R12 Anoctamin 5-related. The distributions of data are represented as histograms along the diagonal. All correlations are significant at the p = 0.05 level.

**Additional Figure 3:**
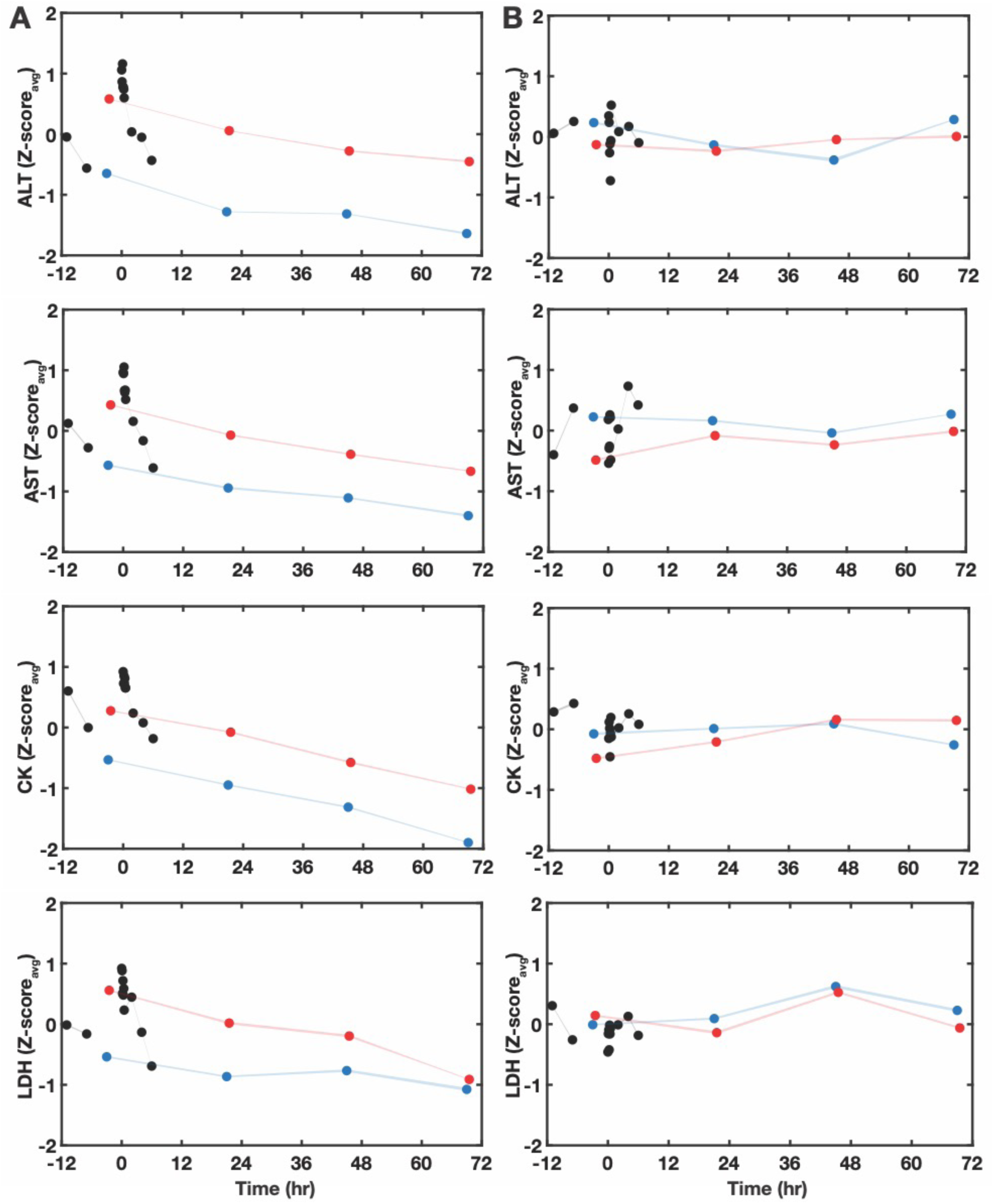
Average Z-score of plasma enzymes with random permutations in time. Average participant plasma ALT, AST, CK, and LDH Z-scores, without (A) and with (B) random permutations in time, plotted against time (in hours) with physical assessment at time 0. Blue and red timepoints correspond to pre- and post-morning activity; black points correspond to post-arrival and post-physical assessment.

**Additional Table 1:** A simplified model was used to fit the log transformed data, with one time constant for the high-frequency data, as the full feature model used for the Z-score data did not converge when applied to the log transformed data.

| *Feature model* | **Fitted Values (-,+ 95% CI)** |  |  |  |
| --- | --- | --- | --- | --- |
|  | *ALT* | *AST* | *CK* | *LDH* |
| Tau 2 (hr) | 41.82 (29.8, 53.85) | 31.3 (23.44, 39.16) | 34.59 (24.36, 44.82) | 36.87 (18.82, 54.92) |
| Tau long (hr) | 691.5 (398.7, 984.2) | 518.6 (326.4, 710.8) | 344 (253.8, 434.2) | 362 (142.7, 581.2) |

